# HPV Vaccination in immunosuppressed patients with established skin warts and non-melanoma skin cancer: A single-institutional cohort study

**DOI:** 10.1101/2023.06.16.23291486

**Authors:** Simon Bossart, Cloé Daneluzzi, Matthias B. Moor, Cédric Hirzel, S. Morteza Seyed Jafari, Robert E. Hunger, Daniel Sidler

## Abstract

**Background:** cSCC (cutaneous squameous cell carcinom) and its precursors are a major cause of morbidity especially in immunosuppressed patients and are frequently associated with human papilloma virus (HPV) infections.

**Objective:** The purpose of this study is to investigate the therapeutically potential of alpha-HPV vaccination for immunosuppressed patients with established cSCC and its precursors.

**Methods:** In this retrospective study, all patients who received Gardasil-9®, a nonavalent HPV vaccine, as secondary prophylaxis were examined. Dermatologic interventions in both the pre- and post-vaccination periods were analyzed with zero-inflated poisson regression and a proportional intensity model for repeated events with consideration of the clinically relevant cofactors.

**Results:** The hazard ratio for major dermatologic interventions was 0.27 (CI 0.14-0.51, p <0.001) between pre- and post Gardasil-9® intervention. Gardasil-9® vaccination showed good efficacy in reducing major dermatologic interventions even after correction of relevant cofactors and national COVID-19 case loads during the observational period.

**Limitation:** The retrospective study design and the rather low number of patients may influence study results. Furthermore, analysis of HPV types and data collection on vaccine-specific HPV antibody measurements was not possible.

**Conclusion:** Alpha-HPV vaccination may potentially cause a significant decrease in dermatologic interventions in immunosuppressed patients with high skin tumor burden.

**Capsule summary:**

- Little is known about a possible immunizing effect of alpha-vaccines in immunosuppressed patients with high skin tumor burden
- Alpha-HPV vaccination such as Gardasil-9® may potentially cause a significant decrease in dermatologic interventions in IS patients with high skin tumor burden.

## Introduction

Skin warts and especially non-melanoma skin cancer (NMSC) are a major cause of morbidity in patients under chronic immunosuppressive treatment, such as patients after solid organ transplantation.^1-2^ The incidence of NMSC increases with the duration of immunosuppressive therapy and develops mainly in sun-exposed areas. While cutaneous squamous cell carcinoma (cSCC) is the most common skin cancer in transplant recipients, occurring 65-250 times more frequently than in the normal population, basal cell carcinoma is increased by a factor of 10 after organ transplantation. ^2-3^ In addition, the occurrence of NMSC is associated with the occurrence of multiple keratotic lesions such as warts and premalignant actinic keratoses as well as intraepithelial neoplasia (Bowen’s disease), which can occur mainly on sun-exposed keratinised skin but also on mucous membranes. ^3-4^ Human Papilloma Virus (HPV) appears to play an important co-carcinogenic role. The incidence of squamous cell carcinoma in transplant recipients increases with the number of keratotic lesions with detection of HPV DNA in 65-90 % of cases. The tumours also frequently show histological features of HPV infection and are usually difficult to distinguish clinically from normal skin warts.^5-7^

HPV infections, which belong to the DNA viruses, affect the keratinocytes of the skin and mucosa. HPVs that affect the mucosa are classified as alpha HPVs. These are divided into low-risk types such as HPV 6 and 11, which induce most condylomas, and high-risk types such as HPV 16 and 18, which are involved in carcinogenesis of the uterus, cervix and orogenital pharynx. HPVs that affect keratinised skin include the alpha-, beta-, gamma-, Mu- and Nu-HPV genera. ^8^

Beta-HPV infection in keratinised skin appears to play a predisposing role in the genesis of NMSC and has been significantly clustered in cSCCs of immunosuppressed transplant recipients. The carcinogenic effect is seen as a combination of beta-HPV infection with ultraviolet (UV) light. ^9-10^

As prevention of HPV infection, only vaccines for alpha-HPV infection have been developed and licensed. Vaccines for the prevention of beta-HPV strains are currently still under investigation.^10^ The three licensed HPV vaccines (Cervarix®, Gardasil®, and Gardasil-9®) on the market are designed to prevent α-HPV related diseases and provide highly effective protection specific to oral and anogenital mucosal HPV subtypes. ^10-11^ The target of this vaccine is the viral capsid L1 of specific alpha HPV strains in mucosal types. Alpha-HPV and beta-HPV, however, show similarities in the immunogenic L1 and L2 capsid proteins.^10,12^ In this regard, there is anecdotic evidence that HPV vaccination against alpha-genera has also resulted in regression of cutaneous warts and cutaneous SCCs. ^13-16^ The similar expression of capsid proteins within the alpha and beta HPV genera could be a possible explanation in this regard. Also, mixed infections of alpha and beta HPV are discussed, explaining the response of vaccination against alpha strains.^10,12-16^

However, no definitive explanations or systematic studies exist to investigate a therapeutic potential of alpha-HPV vaccination for NMSC or its precursors and warts.

This work evaluates recent activity of HPV immunization in immunosuppressed organ transplant recipients (and patients) with recurrent skin warts and non-melanoma skin cancer. In this regard, it must also be noted that patients with severe morbidities payed a relevant toll during the COVID-19 pandemic, and this is specifically true for patients with solid organ transplantations and/or immunosuppressive therapies.^17^ These patients require frequent screening examinations, including dermatology visits to reduce NMSC burden. Therefore, any retrospective analysis of treatment interventions in this field should be anlaysed with consideration of local COVID-19 caseloads and regulatory interventions.^18^

## Methods

### Study Design and population

This study is a registry analysis of patients treated with HPV vaccine Gardasil-9® for the secondary prophylaxis of recurrent keratinotic skin lesions at the University Hospital Inselspital Bern at the Department of Nephrology and Dermatology. All patients had given informed consent for participation in a prospective registry for solid-organ transplantation, and the study was performed with adherence to the principles of the Declaration of Helsinki. All patients received 3 doses of Garasil-9. Patients were regularly followed and treated for skin lesions based on clinical indication by the treating physician. Visits between 1 year before the first vaccination and 1 year after the last vaccination were analysed. Patients’ characteristics were collected from electronic health records and laboratory analyses extracted via Insel Data Science Center (IDSC) and summarised in Table 1. The study protocol was approved by the local ethical committee (ID 2017-01267).

**Table 1:** Baseline characteristics for patients.

| **Patient Characteristics** |  |
| --- | --- |
| **N = 38** |  |
| **Age (years)** | 62 (50, 73) |
| **Sex (male)** | 28 (76%) |
| **Previous NMSC tumor** | 21 (55%) |
| **previous SCC tumors** | 15 (39%) |
| **Only Wart lesions** | 17 (45%) |
| **Underlying disease** |  |
| TPL | 25 (66%) |
| Tumor | 6 (16%) |
| autoimmune | 2 (5.3%) |
| dermatological | 5 (13%) |
| **number of IS** |  |
| 0 | 8 (21%) |
| 1 | 7 (18%) |
| 2 | 10 (26%) |
| 3 | 13 (34%) |
| **localisation** |  |
| solitary | 16 (42%) |
| multiple | 22 (58%) |

TPL: Transplantation

#### Endpoints and co-variates

Primary endpoint of the study was the difference of major dermatological interventions in the pre-vaccination period (1 year before until first vaccination) compared to the post-vaccination period (1 month after third vaccination until end of study). Major dermatological interventions were defined as lesion biopsies, punch biopsies, curettages and excisions. Secondary endpoints were difference of minor interventions (topical treatment and cryotherapies) and overall dermatological visits in the pre- vs. post-vaccination period. An additional endpoint was a logistic regression analysis of vaccination response adjusted for baseline risk factors (frequency of pre-vaccination interventions, age at vaccination, number of immunsuppressive drugs, indication for immunosuppression (IS) [transplant vs. non-transplant]) for recurrent skin tumors.

### Statistical analysis

Results were reported as number of participants (percentage) for categorical data and median (interquartile range) for continuous data. Results were expressed as multivariable-adjusted mean ± SD for categorical values and as adjusted regression coefficient for continuous variables. A two-tailed p<0.05 was considered statistically significant. The primary and secondary endpoints were analysed using a zero-inflated poisson regression. The PWP gap time model was used with a cox proportional hazard model using patient_id as cluster variable and event incidience as strata variable.^19^ The PWP model was analysed in three ways: a crude model with time to event as sole independent predictor, an intermediate model with patients age, gender, type of immunosuppression and type of skin tumor as potentially co-founding factors and a full model with national wide COVID-19 prevalence as additional cofounding factor to account for reduced activity in non-vital medical care during the COVID-pandemic.^17-18^ Statistical analyses were performed using R (version 4.0.3) using tidy verse packages for data analysis and visualisation and survival and pscl for modelling.

## Results

38 patients who received three doses of the nonavalent HPV vaccine Gardasil-9® between December 5th, 2018 and January 1st, 2022 due to NMSC and/or recurrent skin warts were identified and included in the study. 76% of patients were male, mean age was 62 years (IQR: 50, 73). 25 (66%) were treated for SOT, 6 (15%) for tumors and 2 (5%) for autoimmunity and the remainder 5 (13%) for recalcitrant warts. 8 (21%) patients were without immunosuppression, 7 (18%) under mono-, 10 (26%) under dual and 13 (34%) under triple therapy. Among all patients, 16 (42%) received a CNI, 18 (47%) an antimetabolite (Azathioprine or Mycophenolate), 6 (16%) an mTor inhibitor, 17 (45%) PDN and 9 (24%) other IS (supplementary table 1).

Before study begin, 21 (55%) patients were diagnosed with NMSC (15 SCC, 6 CIS) and 17 (45%) with warts without NMSC. 16 (42%) patients had had solitary lesions, 22 (58%) patients multiple lesions at sunexposed localisations (acral, facial). Patient characteristics are summarized in Table 1. All patients received 3 Gardasil-9® vaccinations according to a 3-dose schedule with a median interval of 1.8 months (IQR 1.7-2.1 between first and second and 6.0 months (IQR 5.6-6.1) between first and last months.

Overall 1040 visits with 915 interventions (422 minors and 493 majors) in 100.0 patient years were analysed. 318 visits and 285 interventions (102 minor, 183 major) between first vaccination and one month after last vaccination were included from the analysis. Major interventions included biopsies (n=69), curettages (n=538) and excisions (n=79), minor interventions cryotherapies (n=578) and topical treatments (n=882). 171 visits were without interventions. 425 visits in 37 patient years were analyzed before vaccination and 297 visits in 38 patient years after vaccination (Table 2).

**Table 2:**
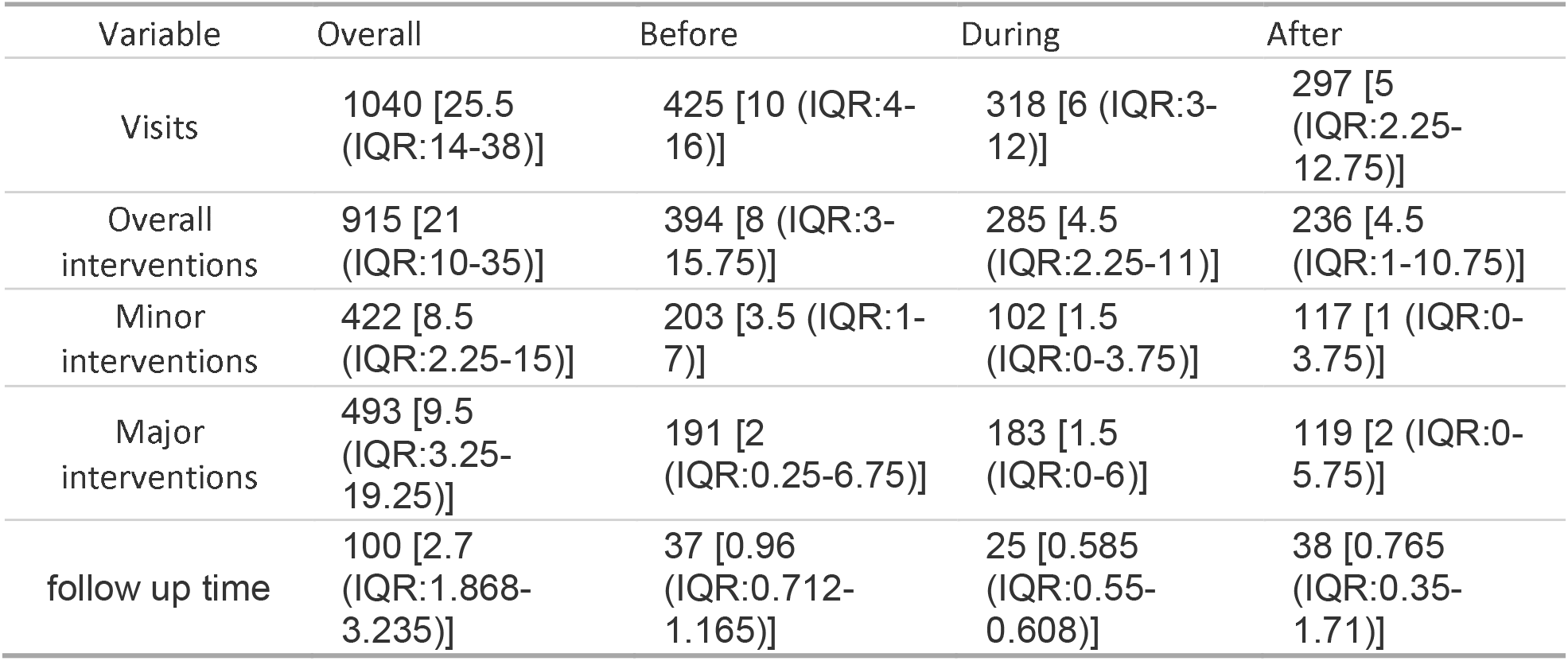
Overview of all patient visits and interventions before, during and after Gardasil-9® vaccinations.

Major dermatological interventions declined after vaccination from a mean of 0.7/year [IQR: 0.0-0.8] to 0.2/year [IQR: 0.0-0.2] (p<0.05) and 68.4% (26/38) of patients experienced a decline in major interventional events. Additionally, minor interventions declined from a mean of 0.6 [IQR: 0.1-0.9] to 0.1/year [IQR: 0.0-0.1] (p<0.01) with a positive response in similar 68.4% of patients (26/38). Similar, overall visits declined from 1.3 to 0.4/year (p<0.01), while visits without any interventions remained stabile (0.1/year to 0.1/year, p=n.s.), see Figure 1.

**Figure 1:**
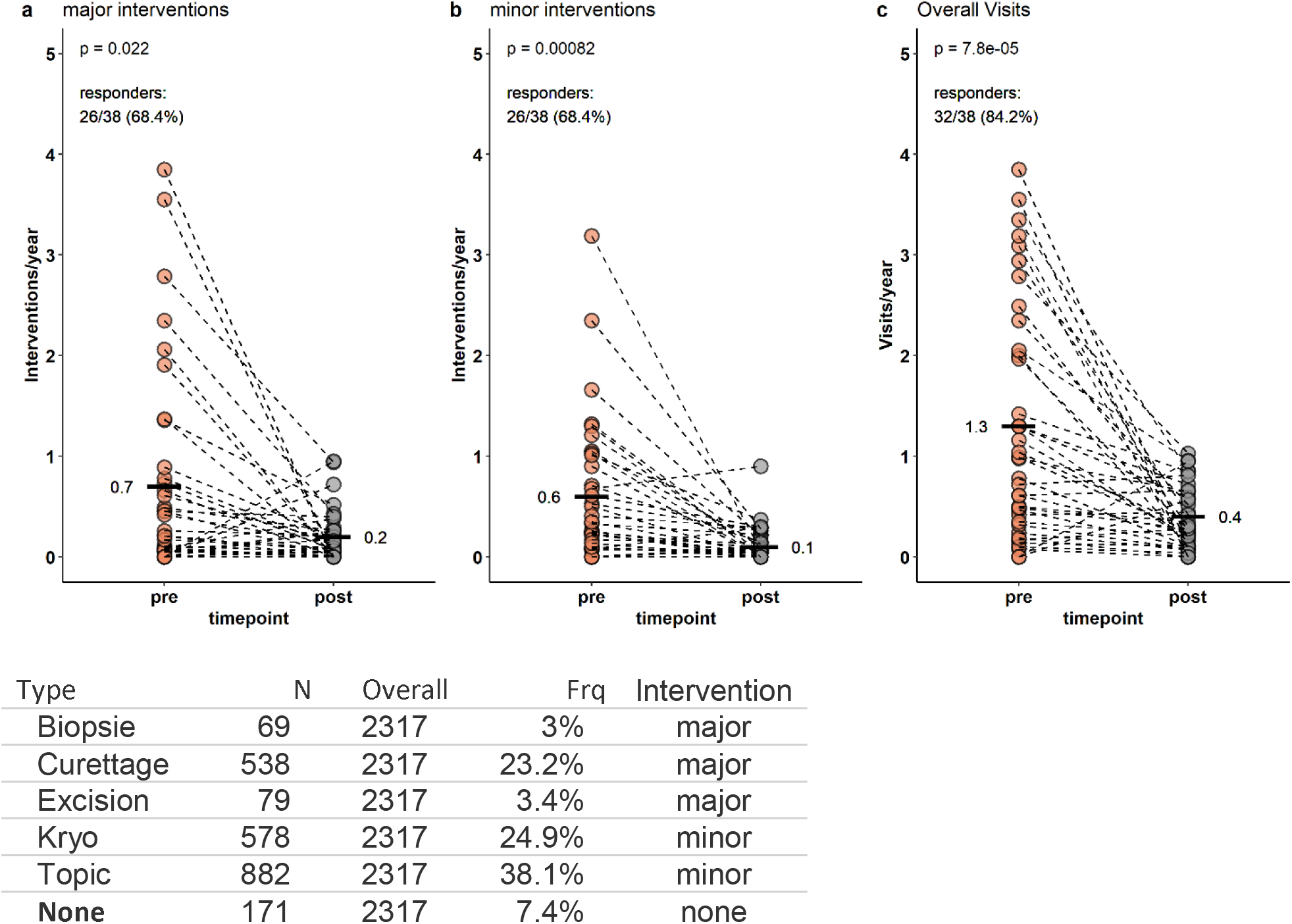
Interventions during study period.

To further these investigations, we performed a time to event-analysis for repeated major dermatological events for each patient. Interestingly, hazard radio for major intervention was 0.27 (CI 0.14-0.51, p <0.001) between pre –and post-Gardasil-9®. Gardasil-9® intervention remained effective in reducing major dermatological interventions after correction for relevant co-factors (full model, HR 0.2, CI 0.11-0.37, p<0.001) and even after correction for national COVID-case load (full model + COVID, HR 0.17, CI 0.09-0.33, p<0.001). Since a relevant proportion of patients received their vaccination schedule during or in the early phase of the COVID pandemic (21.1% completed their vaccination before, 78.9% after start of COVID shutdown in Switzerland on March 16^th^, 2020; Table 3).

**Table 3:** Repeated time to event-analysis for Gardasil-9® Vaccination

|  | crude model |  |  | full model |  |  | full + COVID |  |  |
| --- | --- | --- | --- | --- | --- | --- | --- | --- | --- |
|  | HR<sup>1</sup> | 95% CI<sup>1</sup> | p-value | HR<sup>1</sup> | 95% CI<sup>1</sup> | p-value | HR<sup>1</sup> | 95% CI<sup>1</sup> | p-value |
| **Vaccination** | 0.27 | 0.14,<br>0.51 | **<0.001** | 0.21 | 0.11,<br>0.41 | **<0.001** | 0.19 | 0.08,<br>0.43 | **<0.001** |

<sup>1</sup> HR = Hazard Ratio, CI = Confidence Interval

Dermatological tumor occurrence is dependent on baseline parameters including patient age, history of dermatological interventions, treatment regimen and treatment indication. To study this, we performed logistic regression analysis in a GLM (generalized linear model) with vaccination response (decline of annual major interventions, 26/38 patients) as dependent and previous interventional burden, age at vaccination, number of immunosuppressive drugs and treatment indication (TPL vs. non-TPL) as independent factors. Indeed, TPL patients (OR 7.1 [CI: 0.83-76.8], p=0.086) and heavy pre-vaccination treatment burden (OR 3.68, [CI: 1.06-17.7], p=0.062) was associated with increased odds ratio for treatment response. (Table 4).

**Table 4:** Regression analysis in a GLM with vaccination response

| **GLM for Response** |  |  |  |
| --- | --- | --- | --- |
|  | **OR<sup>1</sup>** | **95% CI<sup>1</sup>** | **p-value** |
| **Frq interventions pre-vacc** | 3.68 | 1.06, 17.7 | 0.062 |
| **age at vacc (years)** | 1.00 | 0.97, 1.03 | >0.9 |
| **no of IS (per 1)** | 0.56 | 0.22, 1.36 | 0.2 |
| **indication (TPL)** | 7.10 | 0.83, 76.8 | 0.086 |

<sup>1</sup> OR = Odds Ratio, CI = Confidence Interval

## Discussion

Cutaneous squamous cell carcinoma and its precursors are the most frequent carcinomas in solid organ transplantation and chronic immunosuppression and correlate with cutaneous human papillomavirus infections, namely the beta-HPV types.^1-4^

The licensed vaccines for HPV immunization are directed against alpha types on mucosa, whereas beta HPV types are mainly found on keratinised skin.^3-4^ To date, there is no good evidence on a possible immunizing effect of these vaccinations against antigens from beta-HPVs on keratinised skin. An effective immunological target of HPVs is the viral capsid envelope, consisting of the minor capsid proteins L1 and L2. Therefore, the currently approved vaccines consist of L1 virus-like particles (VLPs) from certain alpha-HPVs. These vaccines also appear to have cross-reactivity of other alpha-HPV subtypes, but this is highly variable and often low titer.^10^ In addition, cross-reactivity has also been measured in an RCT with induced antibody titers of cutaneous beta-HPV types.^20^ Although this seroconversion of antibodies against cutaneous HPV was rather low in titer and the variability large in the affected individuals, there is anecdotic evidence that clinical regression of cSCC and its precursors can occur after administration of licensed alpha-HPV vaccination.^12-14^

We here report a reduction in dermatology interventional treatment burden in IS patients after administration of an alpha-HPV vaccination, namely Gardasil-9® vaccination. Indeed, median surgical treatment frequency declined by 71% form 0.7/year to 0.2/year within the first 12 months after vaccination. The effect was observed universal throughout various patient groups, involving transplant recipients, older patients and patients with recurrent diseases, but was most experienced in TPL recipients and patients with a previous high tumor burden.

Notably, the recent COVID pandemic with shutdown of general medical services had no impact on interventions during the observation period. Likely, included patients with a high risk of de novo or recurrent skin tumor burden were not restricted to medical limitations.

To our knowledge, this is among the largest cohort of dermatology patients at risk for recurrent keratotic skin lesions investigating the effect of Gardasil-9® vaccination in respect of major surgical intervention. The study population comprised a typical cohort of IS patients in a tertiary dermatology clinic and repeated and well documented treatment log for vaccination and dermatology interventions.

However, this study has some limitations. First the retrospective study design and the rather low number of patients does not allow to exclude all co-founders with may influence study results (patient no-shows, drop-outs). Furthermore, no detailed analysis of HPV infection (type) before and after vaccination and no data on vaccine-specific HPV antibody measurements are available.

The question remains whether a possible seroconversion with mostly low titers against non-genital skin HPV types really has clinical relevance. A possible explanation could be a mixed infection of different HPV types. Thus, sometimes genital mostly high-risk alpha-HPVs are found in cSCC besides beta-HPVs.^21-23^ Thus, antibody generated in L1 vaccines could explain some immunity in skin cancer development in selective cases with high mucosal activity.^10^ However, large-scale prospective RCTs for definitive confirmation are lacking.

Furthermore, attempts are being made to develop vaccination against beta-HPV infections. To date, yet, no anti-beta HPV vaccine is on the market. The large heterogeneity of HPV types in cutaneous keratotic lesions makes the development of efficient anti-beta HPV vaccines difficult.^10^ Also, the direct effect of beta-HPV infection on cutaneous carcinogenesis remains obscure. The development of skin cancer has long been considered as a co-carcinogenic effect of beta-HPV with UV light.^9^ UV irradiation of HPV-infected keratinocytes increases the promoter activity of papillomaviruses and HPV oncoproteins may inhibit the repair of UV-dependent DNA damage in keratinocytes.^9, 24^ In addition, immunosuppressive drugs such as calcineurin inhibitors (e.g. ciclosporin A and tacrolimus) have an inhibitory effect on tumour suppressor genes such as p53, which additionally promotes the development of skin carcinogenesis, and can therefore also have an inhibitory effect on DNA repair mechanisms in the case of UV damage and HPV infection.^9, 25^ Another explanation could be the loss of specific T-cell immunity against the paplloma virus. It has been shown in animal models in rats that suppression of a CD8+T cell-mediated immune response to beta-HPV infection promotes the development of SCCs.^10, 26^ However, these findings do not explain the increased risk of SCC development in immunocompetent patients with beta-HPV infection. A potential explanation could be that in certain immunocompetent patients there is a subclinical dysfunktion of specific CD8+ T-cell or a locally suppression of cutaneous T-cells by UV light on sun exposed sides that favour SCC development.^9-10, 26^

In summary, the association between beta-HPV and cSCC development and the increasing burden of cSCC development especially in immunocompromised patients highlights the need for efficient immunization against HPV-related skin cancer. Although the direct effect of alpha-HPV vaccination in cutaneous HPV infection has not been proven, our study demonstrated a significant decrease in dermatologic procedures in immunosuppressed patients after administration of Gardasil-9® vaccination. Our data show that with administration of such a vaccination, the skin tumor burden in these patients can be significantly decreased. This could reduce invasive procedures and overall mortality as well as healthcare costs. Our data may pave the way for further prospective and randomized controlled trials, which are needed to prove the effect of alpha-HPV in cutaneous HPV infection.

## Data Availability

Raw data are available from the corresponding author on request.

---

**Supplementary table 1:** Immunosuppressive treatments in patients.

| Immunosuppression | n | Frequency |
| --- | --- | --- |
| CNI | 16 | 42% |
| Antimetabolite | 18 | 47% |
| mTor Inh. | 6 | 16% |
| PDN | 17 | 45% |
| Other | 9 | 24% |

CNI: Calcineurin Inhibitor. mTor Inh.: mTor Inhibitor. PDN: Prednisolon

